# Spatiotemporal mapping of major trauma in Victoria, Australia

**DOI:** 10.1101/2021.11.21.21266663

**Authors:** Ben Beck, Andrew Zammit-Mangion, Richard Fry, Karen Smith, Belinda Gabbe

**Author notes:** **Corresponding author:** Dr Ben Beck, School of Public Health and Preventive Medicine, Monash University, Australia, 553 St Kilda Rd, Melbourne, VIC 3004.

## Abstract

**Background:** Spatiotemporal modelling techniques allow one to predict injury across time and space. However, such methods have been underutilised in injury studies. This study demonstrates the use of statistical spatiotemporal modelling in identifying areas of significantly high injury risk, and areas witnessing significantly increasing risk over time.

**Methods:** We performed a retrospective review of hospitalised major trauma patients from the Victorian State Trauma Registry, Australia, between 2007 and 2019. Geographical locations of injury events were mapped to the 79 local government areas (LGAs) in the state. We employed Bayesian spatiotemporal models to quantify spatial and temporal patterns, and analysed the results across a range of geographical remoteness and socioeconomic levels.

**Results:** There were 31,317 major trauma patients included. For major trauma overall, we observed substantial spatial variation in injury incidence and a significant 2.1% increase in injury incidence per year. Area-specific risk of injury by motor vehicle collision was higher in regional areas relative to metropolitan areas, while risk of injury by low fall was higher in metropolitan areas. Significant temporal increases were observed in injury by low fall, and the greatest increases were observed in the most disadvantaged LGAs.

**Conclusions:** These findings can be used to inform injury prevention initiatives, which could be designed to target areas with relatively high injury risk and with significantly increasing injury risk over time. Our finding that the greatest year-on-year increases in injury incidence were observed in the most disadvantaged areas highlights the need for a greater emphasis on reducing inequities in injury.

## INTRODUCTION

Injury surveillance is used to quantify the incidence or risk of injury, identify risk factors, monitor trends over time, identify emerging injury problems, and plan and evaluate injury prevention interventions.^1^ It is well understood that injury incidence is often related to individual-level or neighbourhood-level social, economic and environmental conditions.^2^ While geospatial methods have been widely used in public health to quantify the impact of these area-level factors on disease incidence,^3^ a recent systematic review noted that such approaches have been under-utilised in injury.^4^ Where such methods have been applied, they are often limited to mapping approaches to visually display injury incidence rates and cluster detection methods, and lack the sophistication to robustly quantify and predict spatial variation in injury, and provide measures of uncertainty associated with the predictions.^4^

Amid an increasing availability of geo-referenced administrative data, there has been a growing use of statistical spatial and spatiotemporal models to quantify geographical patterns of disease and how the incidence is changing over time.^3, 5^ Several statistical spatiotemporal models such as the Besag-York-Mollie^6^ model, have been employed to provide estimates of spatial and temporal disease risk. These models can account for spatial autocorrelation (i.e., can account for the fact that areas close together tend to have more similar risk than areas far apart) and draw strength across neighbouring areas to produce more stable estimates of disease risk.^5^ However, such approaches have rarely been applied in the context of injury,^4^ and when used, have been limited to specific causes of injury or nature of injury groups.^7-10^ The general paucity of spatiotemporal models describing serious injury across large geographical areas limits our understanding of how injury risk varies across time and space and our ability to inform geographically targeted injury prevention initiatives. To address this knowledge gap, we aimed to explore spatial and temporal variation of major trauma in Victoria, Australia, through the use of statistical spatiotemporal models. We also used the fitted statistical model to explore the association between regionality and socioeconomic level, and injury incidence.

## METHODS

### Study design

We performed a retrospective review of hospitalised major trauma patients, with a date of injury from 01 January 2007 to 31 December 2019, using data from the Victorian State Trauma Registry (VSTR).

### Setting

The state of Victoria, Australia, has a population of 6.5 million people of which 77% (5 million people) live in the greater metropolitan region of Melbourne. In Australia, local government areas (LGAs) are the third level of government division (beneath states and territories, which are in turn beneath the federal level). There are 79 LGAs in Victoria; 31 are classified as metropolitan and 48 as regional LGAs (Figure 1). In this study, LGAs were selected as the geographical area for analysis. Victoria covers an area of 227,213 km^2^, and the median LGA area is 1,533 km^2^ (range: 9 km^2^– 22,082 km^2^).

**Figure 1:**
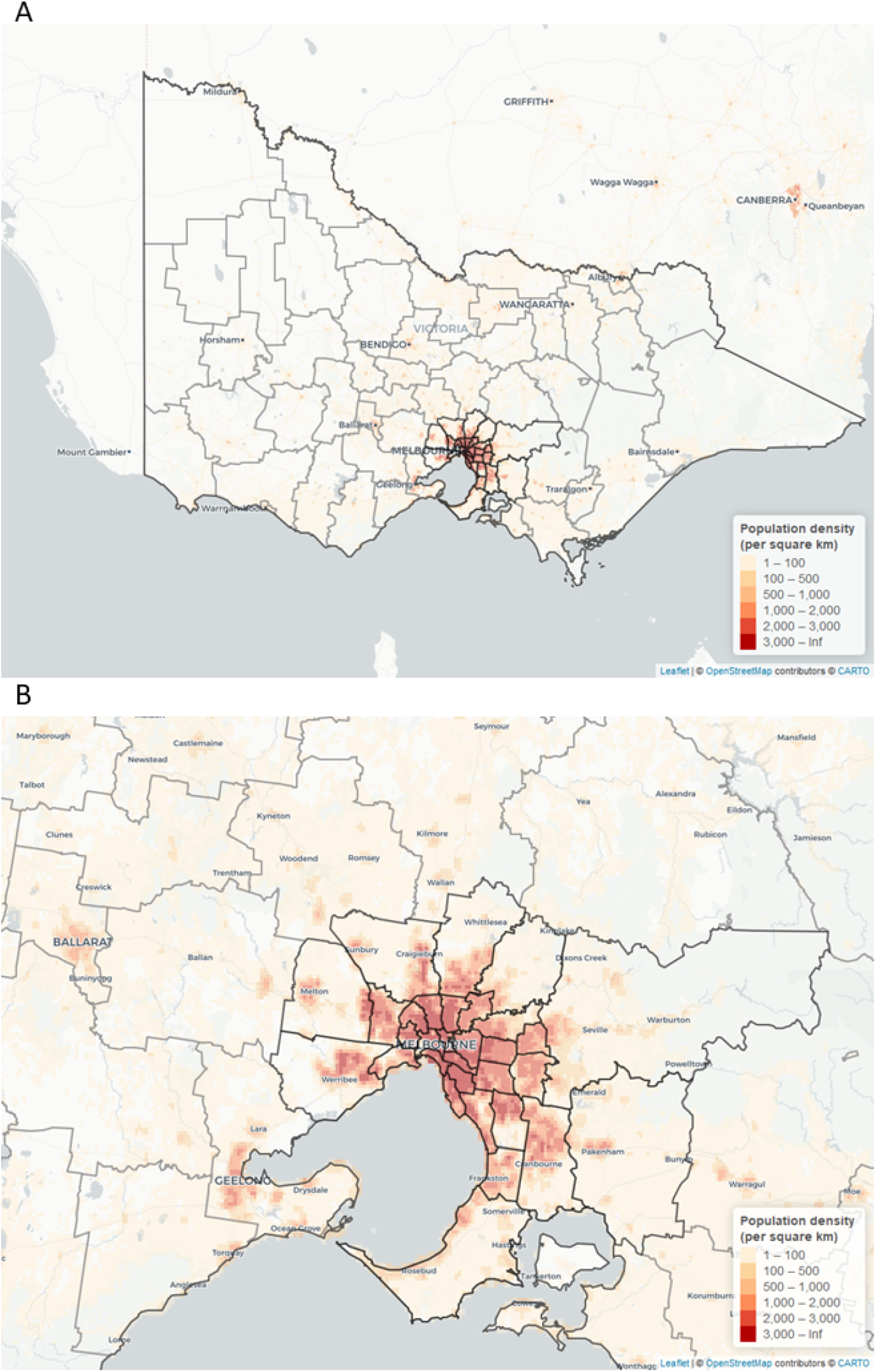
Map of Victoria (A) and Greater Melbourne (B), showing population density overlayed with local government area (LGA) boundaries. Metropolitan LGAs (n=31) are shown with a black border, and regional LGAs (n=48) are shown with a grey border.

### Data sources

The VSTR is a population-based trauma registry that collects data about all hospitalised major trauma patients in the state of Victoria.^11^ A case is included in the VSTR if any of the following criteria are met: (1) death due to injury; (2) an Injury Severity Score (ISS) greater than 12 [Abbreviated Injury Scale (AIS)-2008 update^12^]; (3) admission to an intensive care unit (ICU) for more than 24 hours; and (4) urgent surgery.^13^ Prehospital information is obtained through routine data linkage between the VSTR and Ambulance Victoria (AV). For patients that were transported to hospital by ambulance, precise geographical location (spatial coordinates) of injury events are provided by AV.

### Inclusion criteria

Patients of any age were included in the study if they were transported by ambulance to the primary hospital, had known injury coordinates, and the injury event occurred within the state of Victoria.

### Procedures and statistical analyses

Data on the cause of injury, sex, age and geographical location of the injury event were extracted from the VSTR. Spatial coordinates of injury events were mapped to LGAs. LGA-specific population, remoteness and socio-economic data were sourced from the Australian Bureau of Statistics.^14^ Geographical remoteness of LGAs was measured using the Accessibility and Remoteness Index of Australia (ARIA+). The Victorian Department of Health provides ARIA+ values for each Victorian LGA; these were classified as ‘major cities’, ‘inner regional’ and ‘outer regional’. The socio-economic level of LGAs was measured using the Index of Relative Socio-economic Advantage and Disadvantage (IRSAD), which was classified by the state-level decile, ranging from 1 (most disadvantaged) to 10 (least disadvantaged).^15^ IRSAD summarises information about the economic and social conditions of people and households within an area, and includes measures such as the proportion of individuals with low income; individuals under the age of 70 who have a long-term health condition or disability; individuals with low levels of education; and families with children who live with jobless parents.^15^ As the purpose of our analysis was to quantify the spatial and temporal variation in the incidence of major trauma, we did not seek to explore the association between the different population characteristics and incidence, but rather sought to adjust our estimates for the underlying socio-economic level of each spatial area (LGA). As such, a single comprehensive measure of socio-economic level (IRSAD) was used as a covariate.

We employed Bayesian spatiotemporal models to identify the spatial and temporal patterns of major trauma. These models borrow strength across neighbouring LGAs and time to produce stable estimates of injury risk. Counts of major trauma events in each LGA (*i* = 1, …,79) and year (*t* = −6, …, 6, where t = −6 corresponds to the year 2007 and *t* = 6 to the year 2019) were spatially modelled according to Besag-York-Mollie,^6^ as described elsewhere.^5, 16^ For each area *i* and year *t*, we assume that

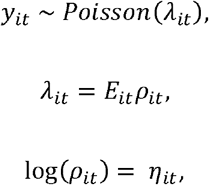

where the number of major trauma cases in region i and year *t, y*_*it*_, is conditionally independent (conditional on *λ*_*it*_) of the number of cases in other regions and years, and Poisson distributed with mean *λ*_*it*_. The corresponding mean number of major trauma cases, *λ*_*it*_, is defined in terms of a risk parameter ρ_*it*_ and the expected number of cases *E*_*it*_, which also varies by region and year. The expected number of cases *E*_*it*_ is set to the overall incidence rate for the entire study area over the study period (2007–2019) multiplied by the LGA-specific population of the corresponding area and year.

Residual variation is captured using the term log(ρ_*it*_) = η_*it*_, for which we consider four models:

1. Model 1: Here, *η*_*it*_ contains a main effect *b*_0_, a spatially structured effect *u*_*i*_, a spatially unstructured effect *v*_*i*_, and a global time trend, where the slope is governed by the parameter *β*:

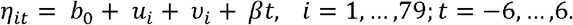
2. Model 2: Here, *η*_*it*_ is as in Model 1, but excludes the spatially structured effect and includes a differential (region-specific) time trend via a space-time interaction term *δ*_i_:

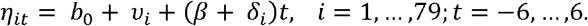
3. Model 3: Here, *η*_*it*_ is as in Model 1, but also contains the differential (region-specific) time trend:

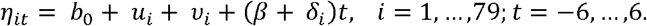
4. Model 4: Here, *η*_*it*_ is as in Model 3, but also uses the socioeconomic level of the LGA (IRSAD; assumed to be time-invariant over the study period) as a covariate with an associated weighting coefficient *a*:

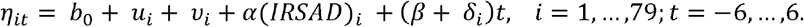

Models 1–4 were fitted using the INLA library^17^ through the statistical software package R v3.6.3 (R Core Team, 2020) and the integrated development environment Rstudio (Rstudio 2020, Boston, MA, USA). The default prior distributions in INLA were employed.

The results from the fitted models are presented through the posterior mean of the area-specific relative risks when compared to the whole of Victoria (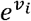 or 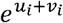, depending on the model), the area-specific yearly multiplicative change in risk (*e*^*β*^ or 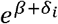, depending on the model), and the posterior mean of the differential time trends (*δ*_*i*_, if present in the model), which reflect the differences between the global temporal trend *β* and the area-specific trends on the linear scale. For the posterior mean of the area-specific relative risk, a value of one indicates that the area-specific risk (averaged over time) is the same as that in the whole of Victoria, a value less than one indicates that it is lower than that in the whole of Victoria, and a value greater than one indicates that it is higher than that in the whole of Victoria. For the posterior mean of the area-specific yearly multiplicative changes in risk, a value of one indicates that the area-specific risk is changing by the same amount year-on-year as that in the rest of Victoria, a value less than one indicates that it is increasing at a lower rate than that in the whole of Victoria, and a value greater than one indicates that it is increasing at a higher rate than that in the whole of Victoria. The posterior probabilities of the area-specific relative risk and yearly multiplicative changes in risk being greater than 1 were calculated.^5^ A comparison of model fit between the four models was done using the Deviance Information Criterion (DIC): a model with a lower DIC is considered to be a better fit to the data than a model with a higher DIC. Three outcomes were modelled independently: the total number of major trauma cases, and the number of major trauma cases for the two leading causes of injury: motor vehicle collisions and low falls (defined as a fall from the same level or from less than one metre). We denote these as the overall, the motor-vehicle collision, and the low fall, studies, respectively.

Comparisons across ARIA+ remoteness classifications and IRSAD socioeconomic quintiles were done through boxplots of the posterior means of the area-specific relative risks and boxplots of the posterior means of the area-specific yearly multiplicative changes in risk.

### Ethical approval

The VSTR has approval from the Department of Health and Human Services Human Research Ethics Committee (HREC) (DHHREC 11/14), the Monash University HREC (CF13/3040 – 2001000165) and 138 trauma-receiving hospitals in Victoria.

Patients or the public were not involved in the design, or conduct, or reporting, or dissemination plans of our research.

## RESULTS

There were 31,317 hospitalised major trauma patients with known injury event coordinates who were included in the analyses (Figure 2). Seventy percent were male, 27% were injured in low fall events and 22% in motor vehicle collisions (Table 1). The overall crude incidence of major trauma was 41.4 per 100,000 population (Figure 3). Histograms of the number of cases in each LGA per year (for major trauma overall, motor vehicle collisions, and low falls) are shown in the Supplementary Material (Figures 1-3).

**Table 1:**
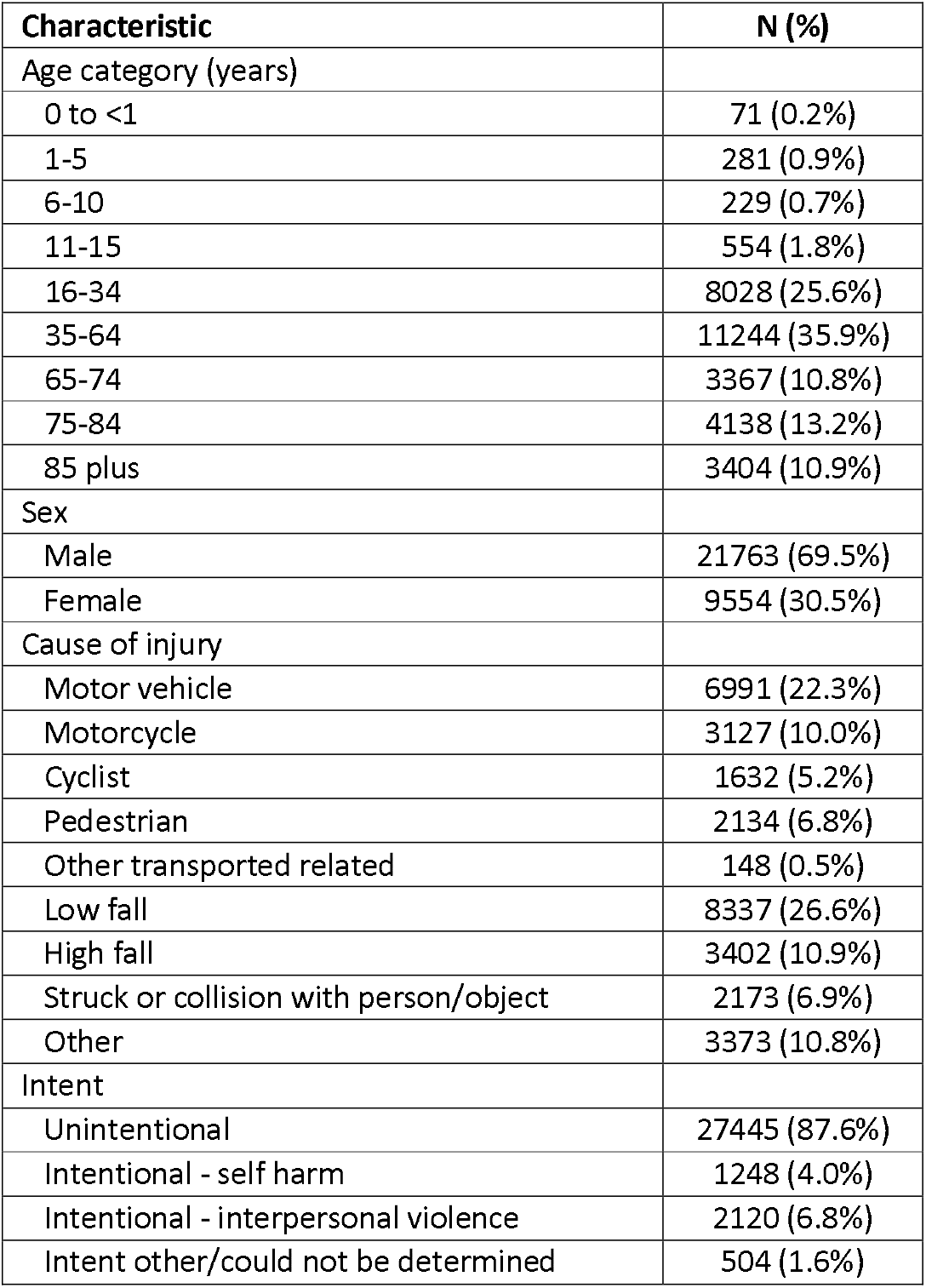
Summary of the data used in our analysis (N=31,317; see also Figure 2).

**Figure 2:**
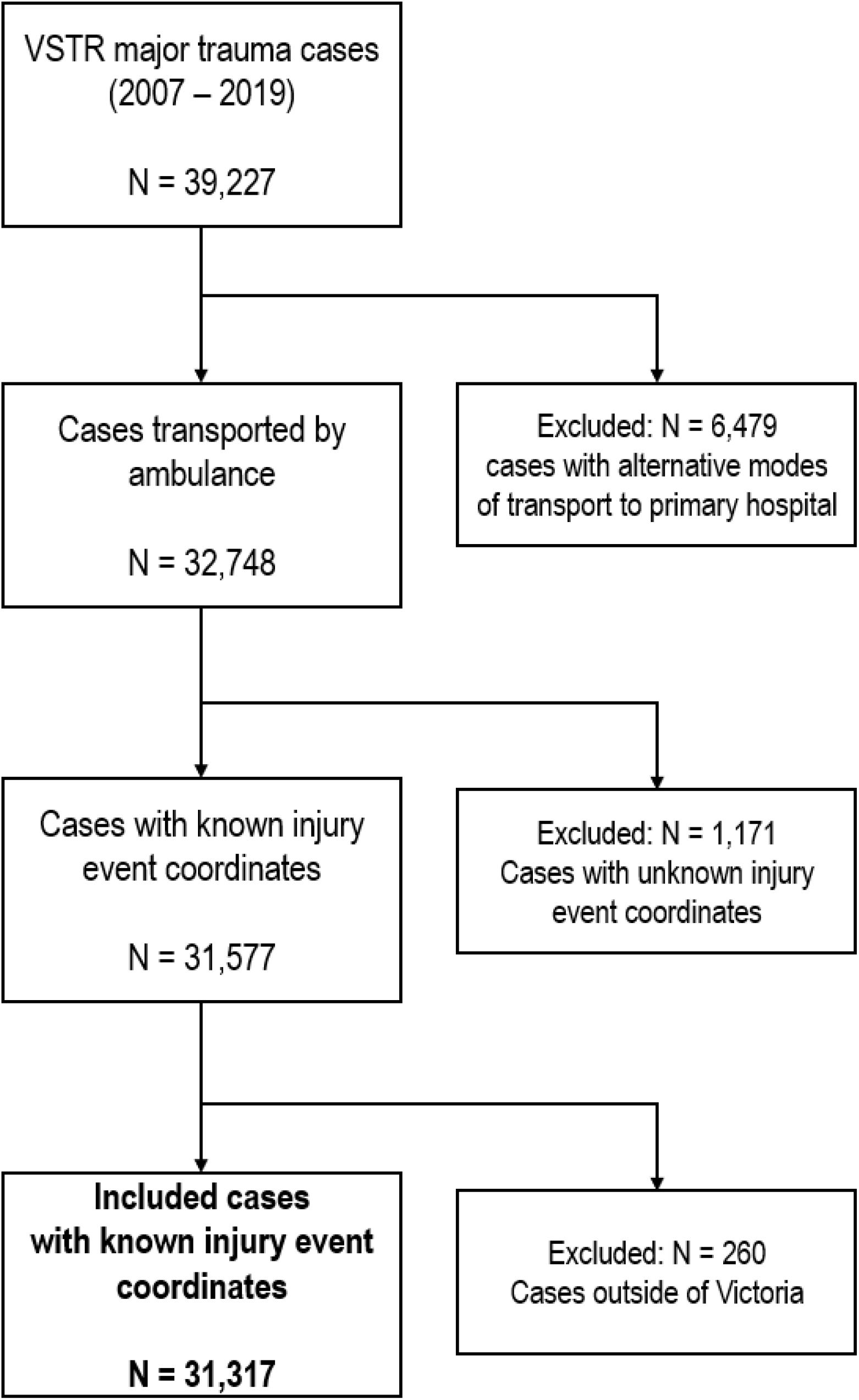
Data-filtering procedure.

**Figure 3:**
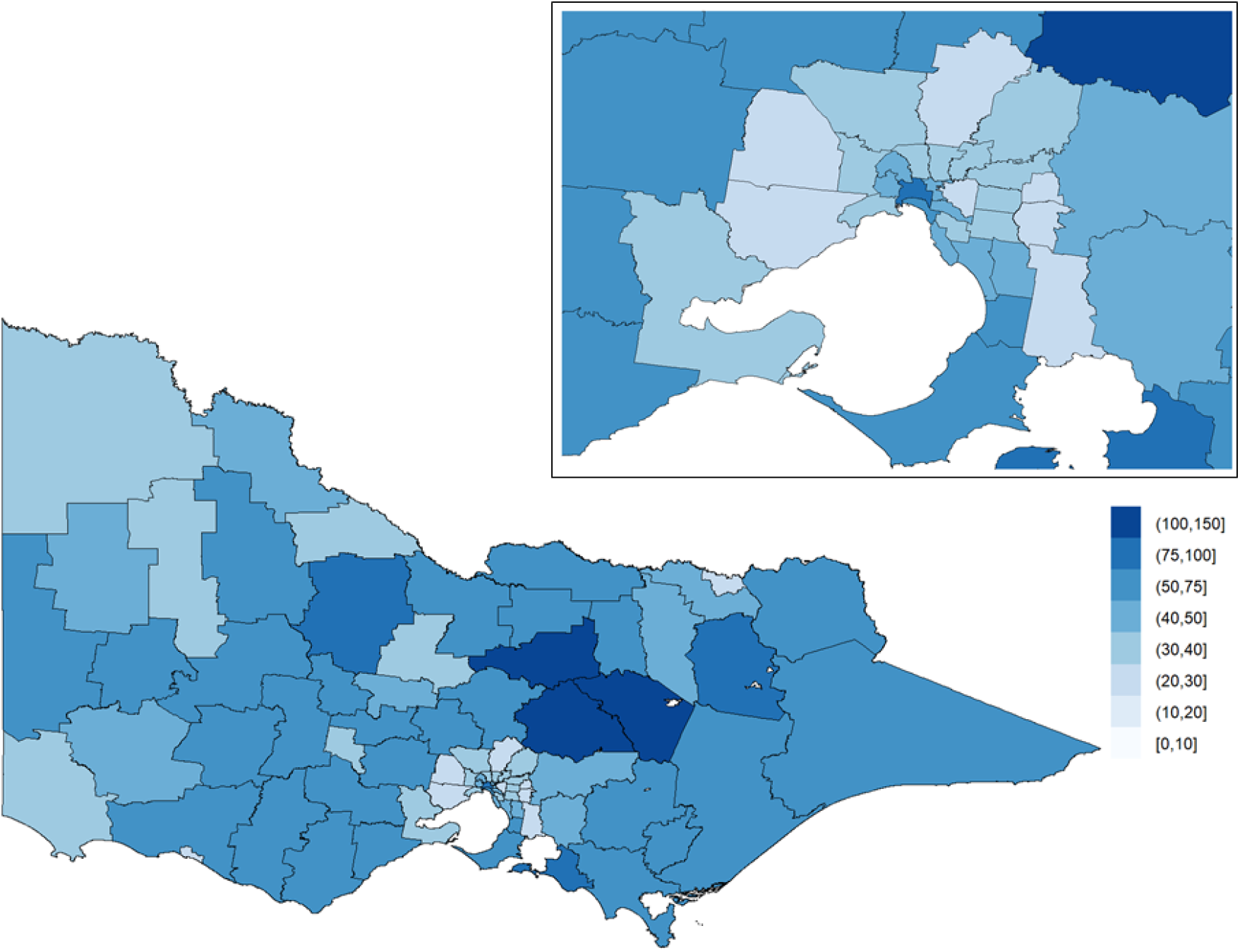
Population-adjusted crude incidence rates of major trauma (per 100,000 population). The inset on the top-right shows Greater Melbourne.

Model fit, in terms of DIC, for the four models, are shown in Table 2. Models with a differential time trend (Models 2,3,4) fit the data substantially better than the model without a differential time trend (Model 1). There was no substantial difference in model fit between Models 2 and 3, and there was also no substantial improvement in model fit with the addition of socioeconomic level as a covariate (Model 4), suggesting that a log-linear relationship between relative risk and socioeconomic status is not appropriate (we empirically explore the association between these two quantities in the results below). The model with the estimated lowest effective number of parameters (Model 3) was chosen as the model with which we proceed with further analysis.

**Table 2:** Model fits for the four models (Models 1-4).

| Model | Deviance Information Criterion (DIC) | Estimated effective number of parameters |
| --- | --- | --- |
| 1) With both a spatially structured effect and a spatially unstructured random effect, but without a differential time trend. | 6461.85 | 75.70 |
| 2) With just a spatially unstructured random effect (independent and identically distributed random variables; IID), and with a differential time trend. | 6259.94 | 122.67 |
| 3) With both a spatially structured effect and a spatially unstructured random effect, and with a differential time trend. | 6259.84 | 122.62 |
| 4) With both a spatially structured effect and a spatially unstructured random effect, with a differential time trend, and with the socioeconomic level of the LGA (measured using IRSAD) as a covariate. | 6259.92 | 122.66 |

### Overall (all major trauma)

The posterior means of the area-specific relative risk for major trauma overall ranged from 0.49 to 2.98 (Figure 4). LGAs with an area-specific relative risk higher than the average for the whole of Victoria were observed in eastern Victoria, northern Victoria and some LGAs in western Victoria. The excess spatial risk, reflecting the probability of an area-specific relative risk being greater than 1, is shown in Supplementary Material (Figure 4). The study-wide yearly multiplicative change in risk over the 13-year study period (i.e., *e*^*β*^) revealed a 2.1% increase in incidence year on year (95% credible interval: [1.7%, 2.5%]). The area-specific yearly multiplicative change in risk is shown in Figure 5. There was no obvious spatial clustering in the area-specific yearly change for major trauma overall, and incidence rate of change across time was mostly positive.

**Figure 4:**
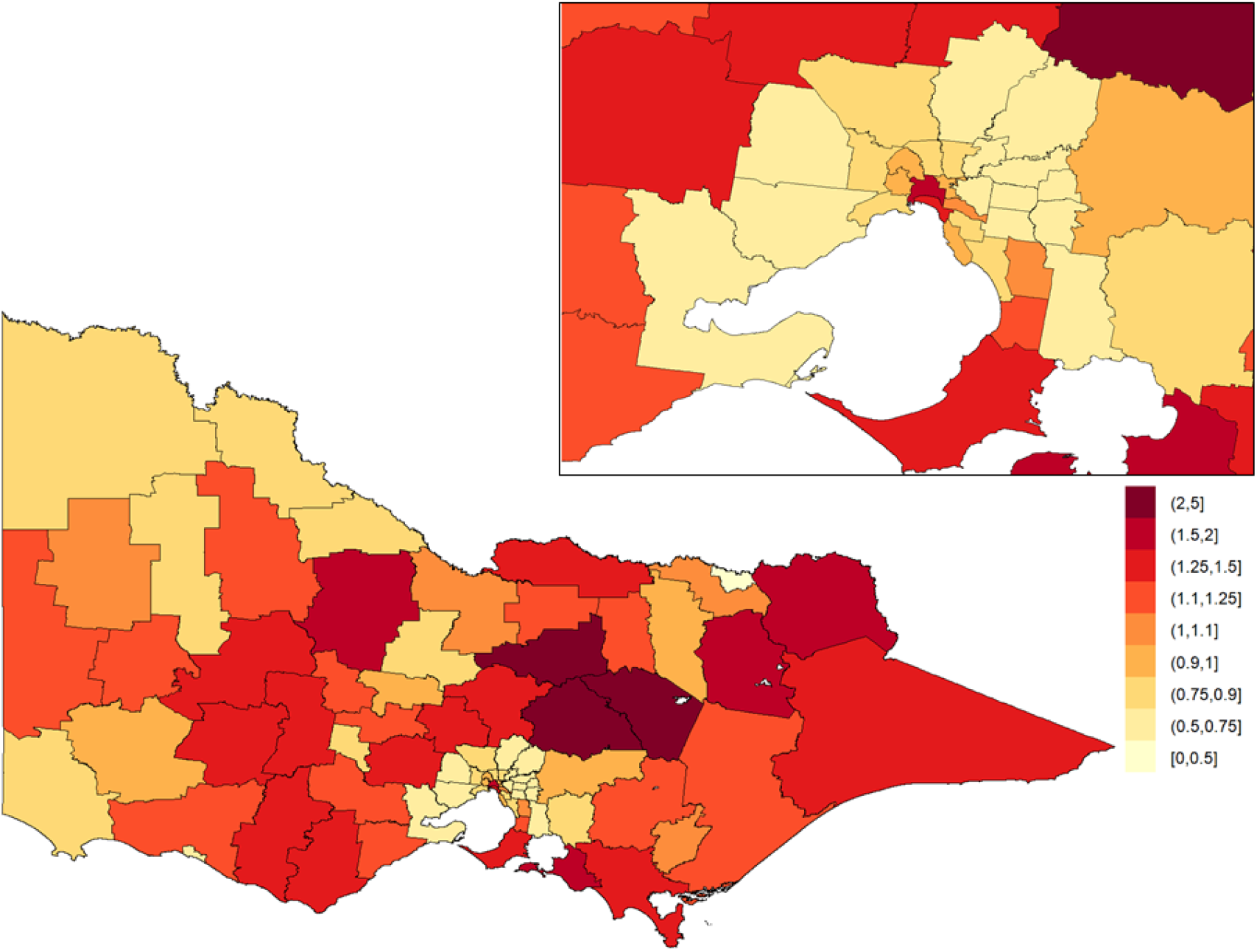
Posterior mean of the area-specific relative risk for major trauma overall in Victoria. Here, a value of one indicates that the area-specific risk (averaged over time) is the same as that in the whole of Victoria, a value less than one indicates that it is lower than that in the whole of Victoria, and a value greater than one indicates that it is higher than that in the whole of Victoria. The inset on the top-right shows Greater Melbourne.

**Figure 5:**
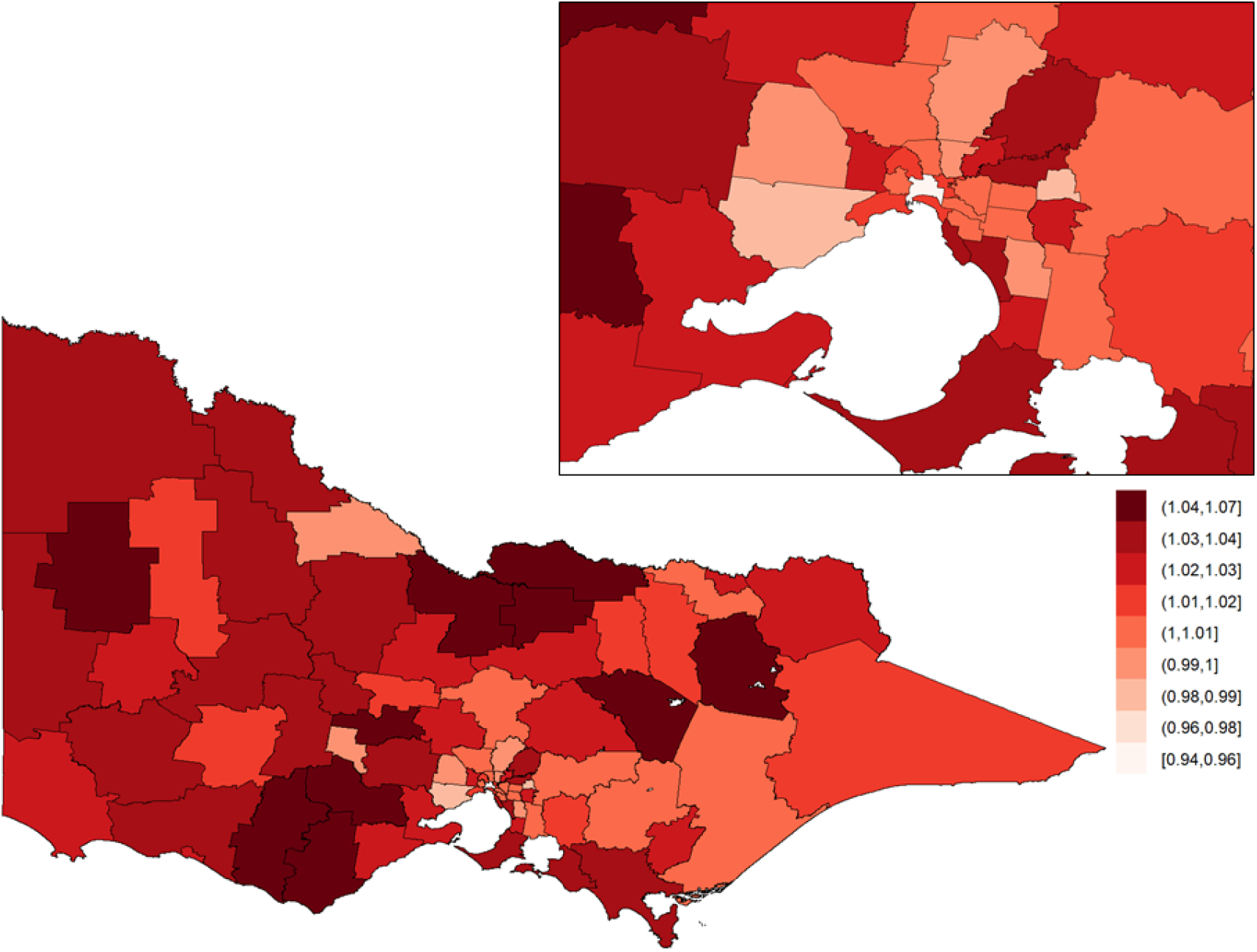
Area-specific yearly multiplicative change in risk for major trauma overall in Victoria. Here, a value of one indicates that the area-specific risk is stable over time; a value greater than one indicates that the area-specific risk is increasing over time; and a value less than one indicates that the area-specific risk is declining over time. The inset on the top-right shows Greater Melbourne.

When contrasting the posterior mean of the area-specific relative risks across ARIA classifications (Figure 10), we find that area-specific relative risks for major trauma overall are higher in inner regional areas (median = 1.10; Q1 = 0.81, Q3 = 1.25, where Q1 and Q3 denote the first and third quartile, respectively) and outer regional areas (median = 1.22; Q1 = 0.95, Q3 = 1.37), when compared to major cities (median = 0.80; Q1 = 0.68, Q3 = 0.97). Positive yearly changes in relative risk were observed across all ARIA classifications; increases were higher in inner and outer regional areas compared to major cities (Table 4).

When contrasting variation across socioeconomic levels (Error! Reference source not found.), we find that area-specific relative risks are higher for more disadvantaged LGAs (IRSAD quintiles 1,2,3) relative to more advantaged LGAs (IRSAD quintiles 4,5) (see also Table 4). With regard to temporal changes, increases over time are slightly higher in more disadvantaged LGAs (IRSAD quintiles 1,2,3) compared to more advantaged LGAs (IRSAD quintiles 4,5) (Table 4).

Posterior probabilities of the spatial risks and the yearly multiplicative changes in risks being greater than 1 for major trauma overall are shown in the Supplementary Material (Figures 4 and 10, respectively).

### Motor vehicle collisions

For motor vehicle collisions, the posterior means of the area-specific relative risk ranged from 0.29 to 4.18, and the area-specific relative risk was higher in regional areas compared to metropolitan areas (Figure 6). The overall 13-year yearly multiplicative change in risk for motor vehicle collisions was not significantly different from 1.

**Figure 6:**
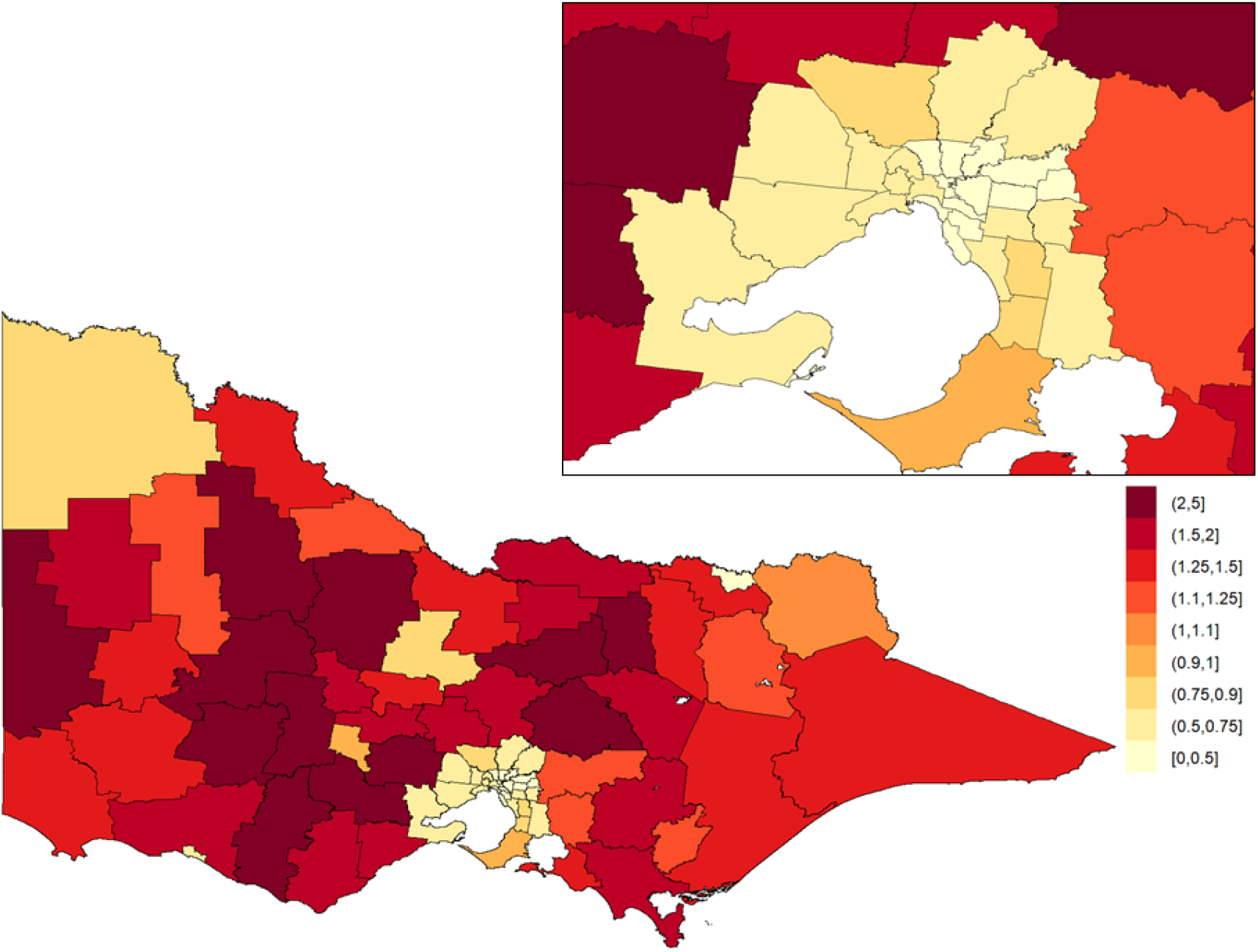
Posterior mean of the area-specific relative risk of injury by motor vehicle collision in Victoria. Here, a value of one indicates that the area-specific risk (averaged over time) is the same as that in the whole of Victoria, a value less than one indicates that it is lower than that in the whole of Victoria, and a value greater than one indicates that it is higher than that in the whole of Victoria. The inset on the top-right shows Greater Melbourne.

**Figure 7:**
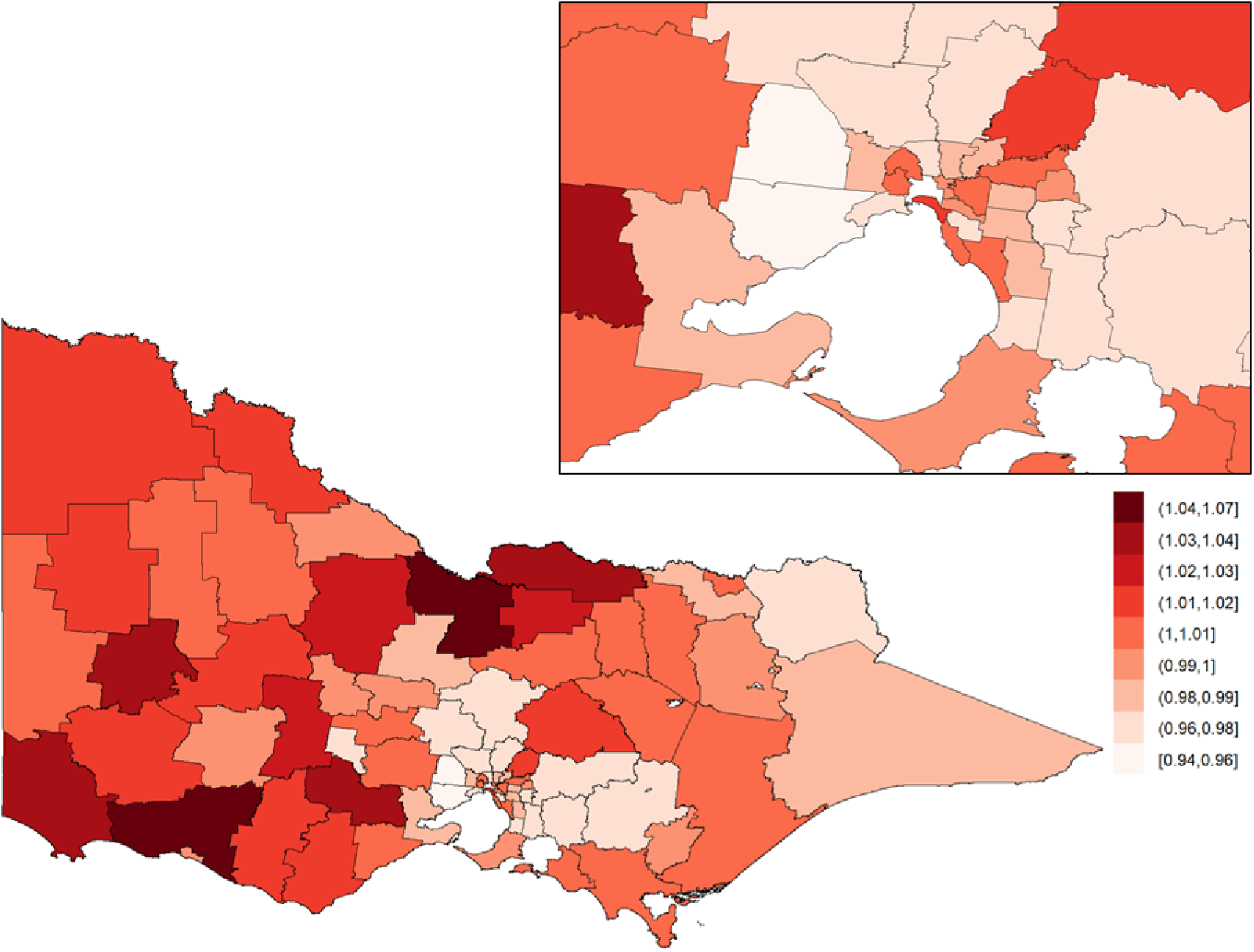
Area-specific yearly multiplicative change in risk of injury by motor vehicle collision in Victoria. Here, a value of one indicates that the area-specific risk is stable over time; a value greater than one indicates that the area-specific risk is increasing over time; and a value less than one indicates that the area-specific risk is declining over time. The inset on the top-right shows Greater Melbourne.

Area-specific relative risks for motor vehicle collisions were consistently higher in inner regional areas (median = 1.48; Q1 = 0.86, Q3 = 1.88) and outer regional areas (median = 1.48; Q1 = 1.30, Q3 = 2.08) when compared to major cities (median = 0.52; Q1 = 0.40, Q3 = 0.63). With regard to temporal trends (Figure 10), an increasing rate of motor vehicle collisions was observed in outer regional LGAs (median = 1.010; Q1 = 1.003, Q3 = 1.016) while decreases were observed in major cities (median = 0.986; Q1 = 0.976, Q3 = 1.001).

Across socioeconomic levels, the area-specific relative risks for motor vehicle collisions were substantially higher in the most disadvantaged LGAs (median = 1.44; Q1 = 1.12, Q3 = 1.99) relative to the most advantaged LGAs (median = 0.43; Q1 = 0.37, Q3 = 0.63). No changes over time were observed in more disadvantaged LGAs (IRSAD quintiles 1,2) while decreases over time were observed in more advantaged LGAs (IRSAD quintiles 3,4,5) (Table 4).

Posterior probabilities of the spatial risks and yearly multiplicative changes in risks being greater than 1 for motor vehicle collisions are shown in the Supplementary Material (Figures 5 and 11, respectively).

### Low falls

For low falls, the posterior means of the area-specific relative risk ranged from 0.51 to 2.57, with the area-specific relative risk higher in metropolitan areas compared to regional areas (Figure 8). The study-wide yearly multiplicative change in risk revealed a 4.0% incidence increase year on year (95% credible interval: [3.3%, 4.8%]). The area-specific yearly change in relative risk was consistently high across all LGAs (Figure 9).

**Figure 8:**
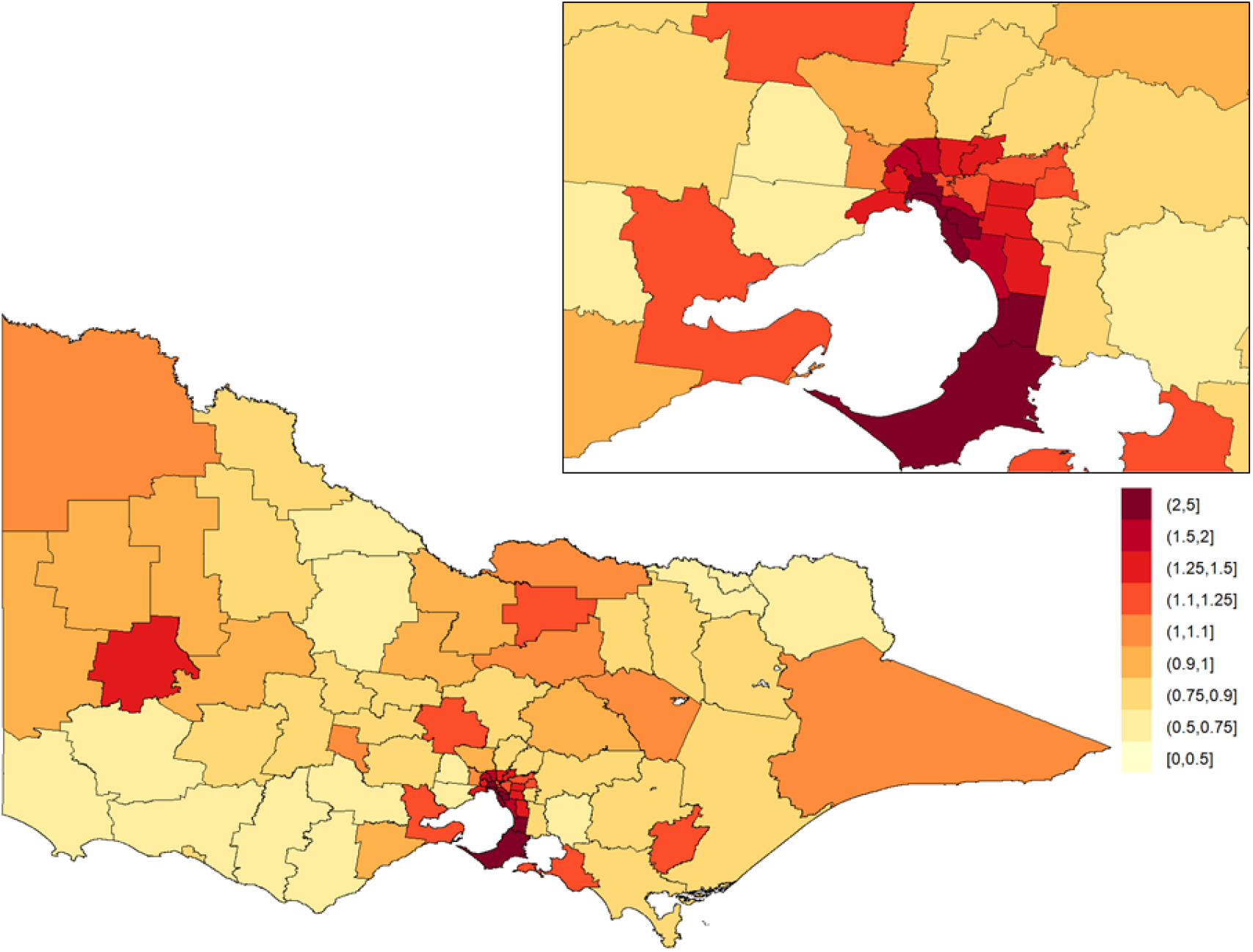
Posterior mean of the area-specific relative risk of injury by low fall in Victoria. Here, a value of one indicates that the area-specific risk (averaged over time) is the same as that in the whole of Victoria, a value less than one indicates that it is lower than that in the whole of Victoria, and a value greater than one indicates that it is higher than that in the whole of Victoria. The inset on the top-right shows Greater Melbourne.

**Figure 9:**
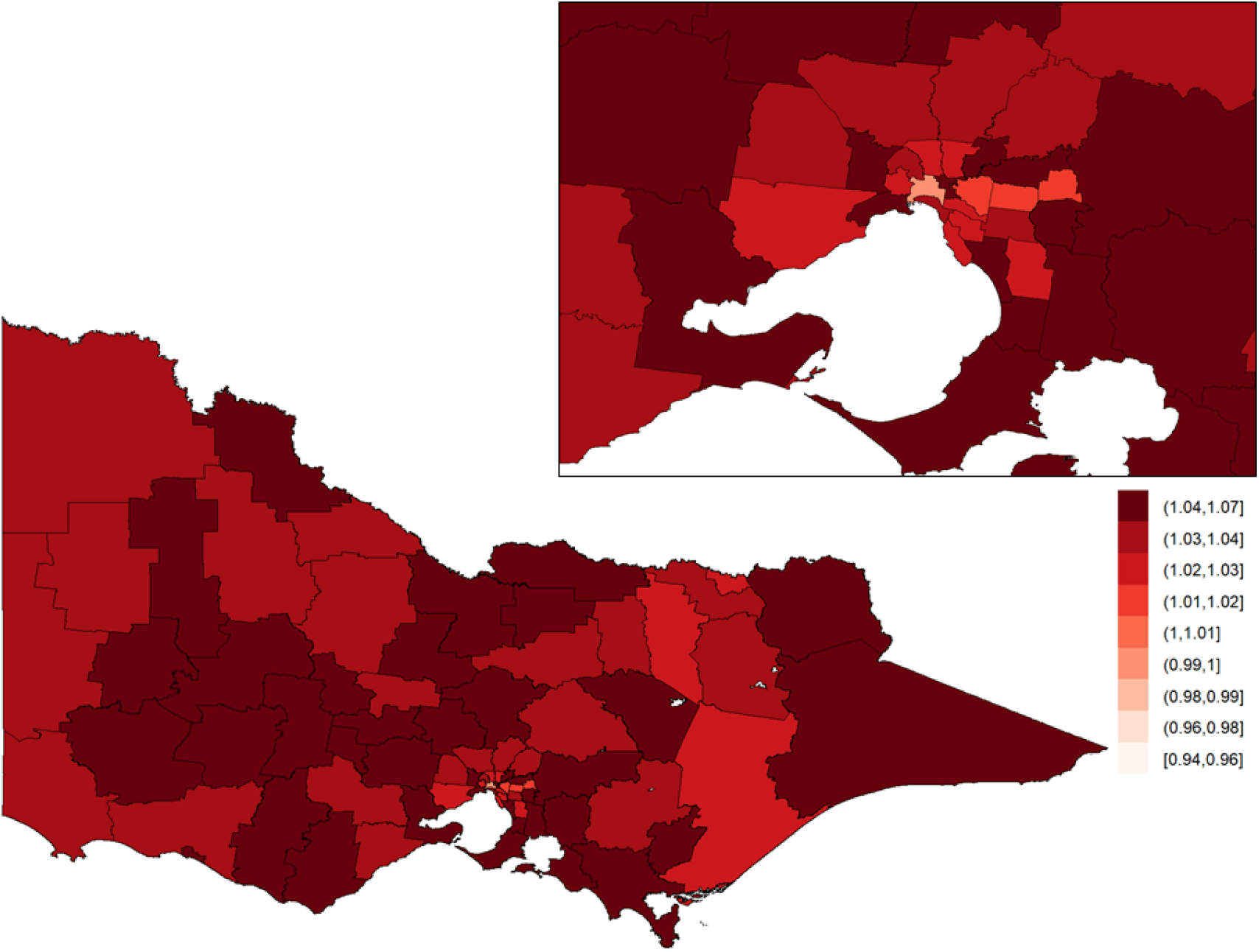
Area-specific yearly multiplicative change in risk of injury by law fall in Victoria. Here, a value of one indicates that the area-specific risk is stable over time; a value greater than one indicates that the area-specific risk is increasing over time; and a value less than one indicates that the area-specific risk is declining over time. The inset on the top-right shows Greater Melbourne.

Area-specific relative risks for low falls were higher in major cities (median = 1.41; Q1 = 1.22, Q3 = 1.91) than in inner (median = 0.87; Q1 = 0.80, Q3 = 1.02) and outer regional (median = 0.82; Q1 = 0.68, Q3 = 0.98) LGAs. Temporal increases in low falls were consistently high across all ARIA classifications (Figure 10, Table 3).

**Figure 10:**
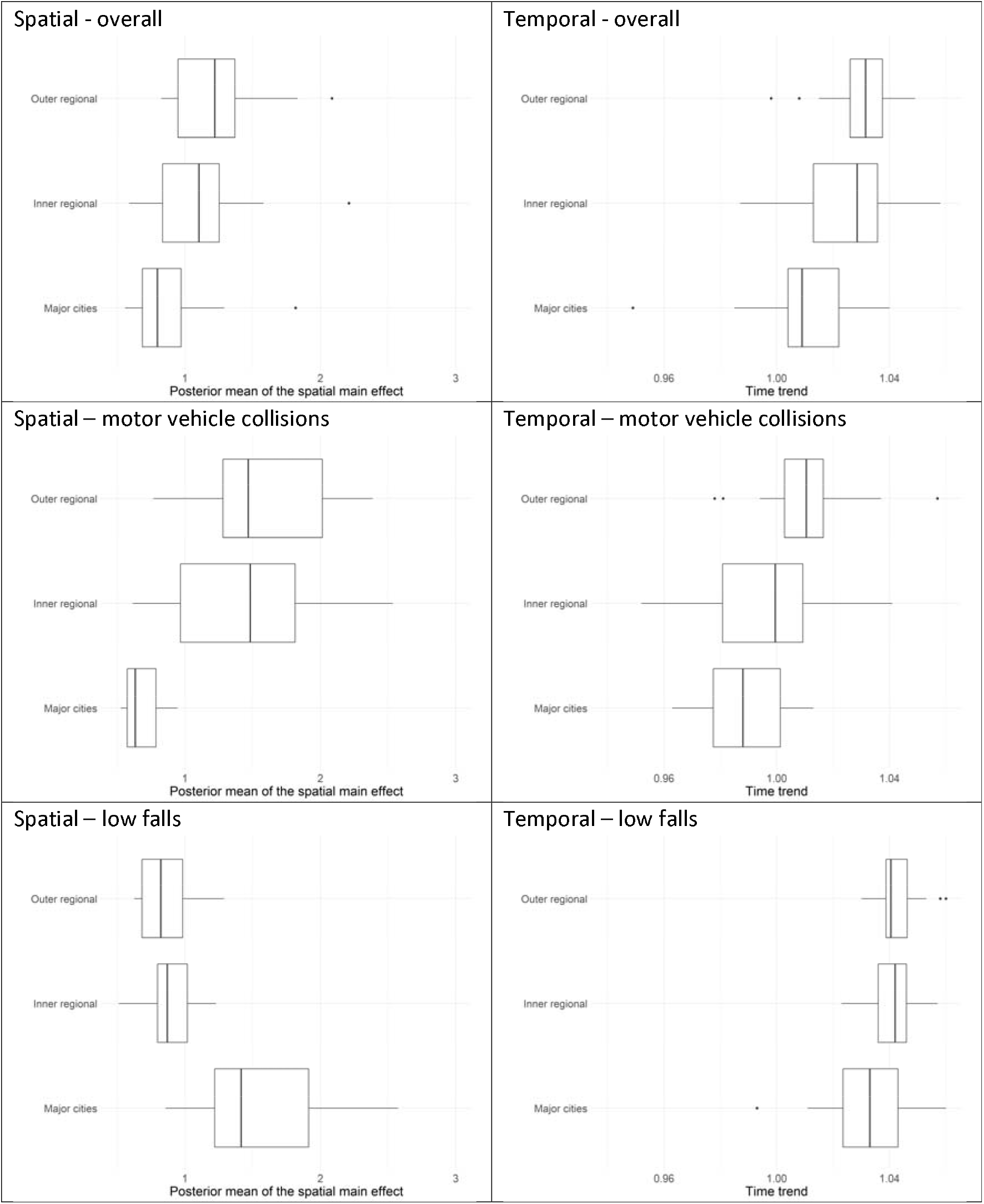
Box plots of posterior means of area-specific relative risk (‘Spatial’) and area-specific yearly multiplicative change in risk (‘Temporal’) of major trauma, shown by major trauma overall, motor-vehicle collisions, and low falls, and by the Accessibility and Remoteness Index of Australia (ARIA), which classifies an LGA as a major city, as inner regional, or as outer regional. The middle line of the box represents the median, with the lower and upper box ranges reflecting lower and upper quartile, respectively. The upper/lower whiskers reflect the largest/smallest observation less than or equal to ± 1.5 times the interquartile range.

**Figure 11:**
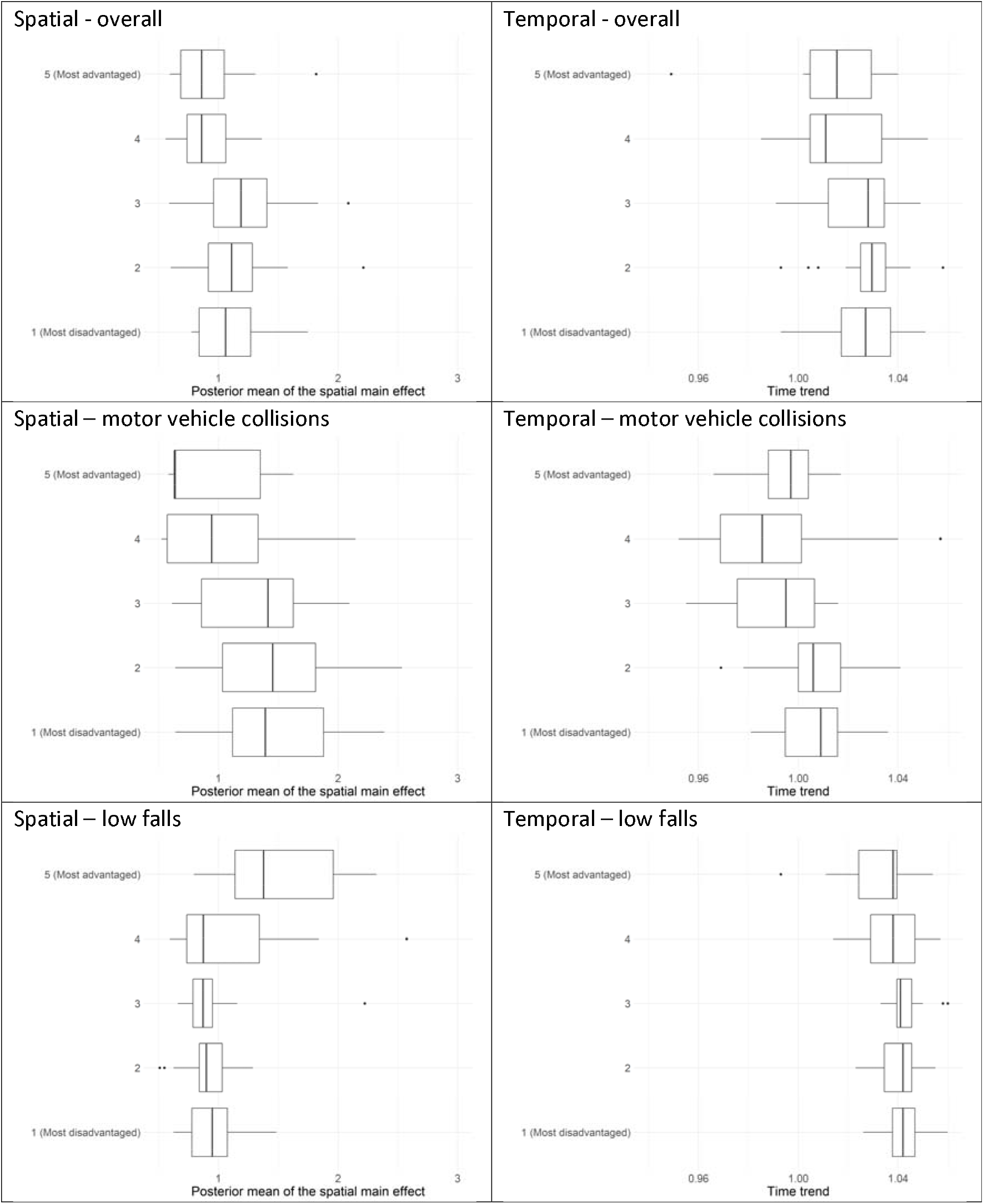
Box plots of posterior means of area-specific relative risk (‘Spatial’) and area-specific yearly multiplicative change in risk (‘Temporal’) of major trauma, shown by study (overall, motor-vehicle collisions, and low falls) and by the Index of Relative Socio-economic Advantage and Disadvantage (IRSAD), which classifies an LGA from 1 (most disadvantage), to 5 (most advantaged). The middle line of the box represents the median, with the lower and upper box ranges reflecting lower and upper quartile, respectively. The upper/lower whiskers reflect the largest/smallest observation less than or equal to ± 1.5 times the interquartile range.

**Table 3:**
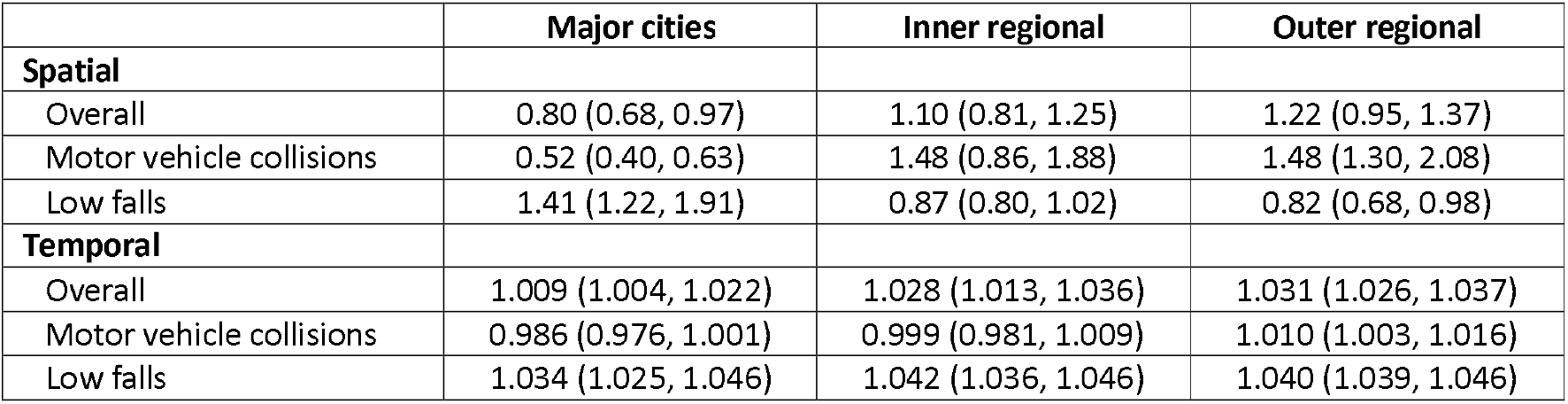
Summaries of the posterior means of the area-specific relative risk (‘Spatial’) and the area-specific yearly multiplicative change in risk (‘Temporal’) grouped by the Accessibility and Remoteness Index of Australia (ARIA), classified as major cities, inner regional and outer regional. Data are presented for major trauma overall, motor vehicle collisions, and low falls. Values represent the median for each group. The first and third quartiles are given in parentheses.

|  | Major cities | Inner regional | Outer regional |
| --- | --- | --- | --- |
| **Spatial** |  |  |  |
| Overall | 0.80 (0.68, 0.97) | 1.10 (0.81, 1.25) | 1.22 (0.95, 1.37) |
| Motor vehicle collisions | 0.52 (0.40, 0.63) | 1.48 (0.86, 1.88) | 1.48 (1.30, 2.08) |
| Low falls | 1.41 (1.22, 1.91) | 0.87 (0.80, 1.02) | 0.82 (0.68, 0.98) |
| **Temporal** |  |  |  |
| Overall | 1.009 (1.004, 1.022) | 1.028 (1.013, 1.036) | 1.031 (1.026, 1.037) |
| Motor vehicle collisions | 0.986 (0.976, 1.001) | 0.999 (0.981, 1.009) | 1.010 (1.003, 1.016) |
| Low falls | 1.034 (1.025, 1.046) | 1.042 (1.036, 1.046) | 1.040 (1.039, 1.046) |

**Table 4:**
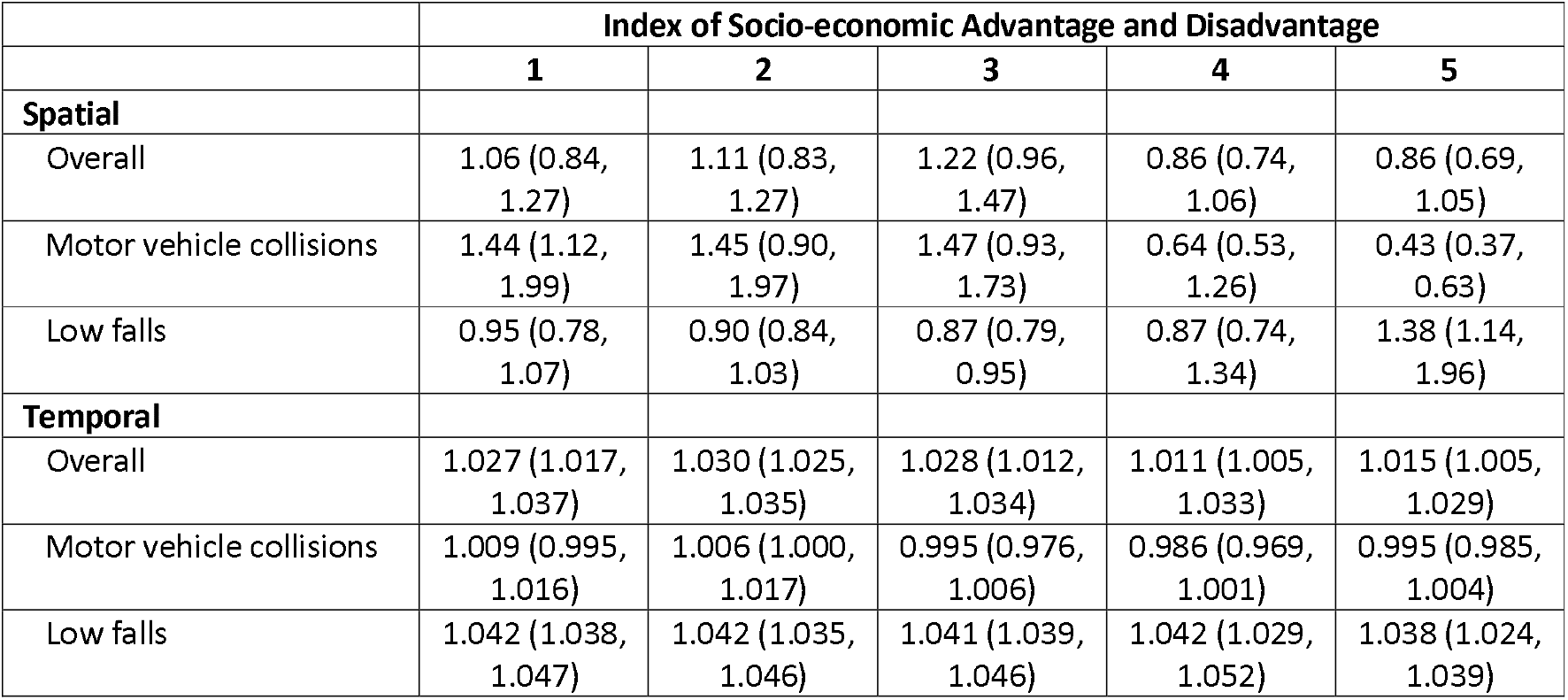
Summaries of the posterior means of the area-specific relative risk (‘Spatial’) and the area specific yearly multiplicative change in risk (‘Temporal’) grouped by the Index of Relative Socio-economic Advantage and Disadvantage (IRSAD), classified from 1 (most disadvantage), to 5 (most advantaged). Data are presented for major trauma overall, motor vehicle collisions, and low falls. Values represent the median for each group. The first and third quartiles are given in parentheses.

Unlike what was observed with motor vehicle collisions, the area-specific relative risks for low falls were lower in the most disadvantaged LGAs (median = 0.95; Q1 = 0.78, Q3 = 1.07) than in the most advantaged LGAs (median = 1.38; Q1 = 1.14, Q3 = 1.96). With regard to temporal trends, increases over time were consistently high across all socioeconomic levels (Error! Reference source not found., Table 4), although the rate of increase was slightly higher in more disadvantaged LGAs.

Posterior probabilities of the spatial risks and the yearly multiplicative changes in risks being greater than 1 for low falls are shown in the Supplementary Material (Figures 6 and 12, respectively).

## DISCUSSION

In this population-based study, we showed through the use of statistical spatiotemporal models that there is substantial, and significant, spatial and temporal variation in the incidence of major trauma. Area-specific injury risk for motor vehicle collisions was higher in regional areas relative to metropolitan areas, while injury risk for low falls was higher in metropolitan areas. Temporal increases were observed in low falls, and the greatest increases were observed in the most disadvantaged LGAs.

To our knowledge, this is the first study to explore spatiotemporal patterns of major trauma via the lens of statistical models. Previous studies employing spatial modelling of major trauma have focused on spatial variation only,^18, 19^cluster analysis,^20^ or only explored access to trauma centres.^21,22^

There are, however, a number of studies that have employed spatiotemporal modelling in other injury cohorts. For example, Jansen *et al*.^23^ used geographically weighted regression to model five years of injury events attended by ambulance in Scotland, demonstrating varying spatial and temporal patterns of injury across small areas. DiMaggio^7^quantified the spatiotemporal risk of pedestrian and bicyclist injury in New York City over a 10-year period, identifying census tracts that were at a persistently high risk of injury. However, this study was reliant on police-reported crashes involving a motor vehicle, which are known to exclude a large number of hospitalised cases that are not reported to police.^24^ However, together with other studies that have used spatiotemporal modelling,^4, 9, 10, 25^ these studies collectively demonstrate the importance of unpacking spatial and temporal trends in small spatial areas.

Our finding that injury rates for motor vehicle collisions were substantially higher in inner and outer regional LGAs than in major cities is consistent with existing Australian and international literature.^26-29^

In Australia, over 70% of the population reside in metropolitan areas, yet over 50% of road deaths occur in regional or remote areas,^29^ and people living in very remote areas are 2.2 times as likely to be hospitalised from a transport crash relative to people living in major cities.^30^ It has been suggested that high speed limits, limited physical separation, unforgiving roadsides, low speed enforcement, and typically poor-quality roads, are contributing factors to higher injury rates in regional areas.^31^

In contrast, low fall injury rates were higher in LGAs classified as major cities than in inner and outer regional LGAs, and higher in the most socioeconomically advantaged LGAs. While low socioeconomic status has been associated with frailty^32, 33^ and functional decline^34^ among older adults, the evidence on the association between socioeconomic level and low fall injury rates is mixed. Some studies have demonstrated an association between high educational level and a decreased history of falls,^35-37^ while others have reported the converse.^38-40^ While we did not calculate age-sex standardised incidence rates to account for regional differences in population distributions, the proportion of older Australian adults living in urban areas is similar to that in regional areas,^41^ and therefore differences in the underlying population do not solely explain the differences we observed between major cities and inner and outer regional areas. Further research is required to understand specific components of socioeconomic and environmental exposures that relate to low fall injury risk.

It is well established that inequities in injury persist within and between populations and environments, and people with lower socioeconomic status are more likely to experience fatal and serious injury.^18, 42, 43^Our findings generally support this, as the median LGA area-specific relative risk was higher in the most disadvantaged LGAs compared to the most advantaged LGAs (median = 1.06 vs 0.86), noting that there was overlap in the interquartile ranges. The effect of a higher area-specific relative risk in the most disadvantaged LGAs relative to the most advantaged LGAs was particularly apparent for motor vehicle collisions (median LGA spatial effect for the most disadvantaged LGAs = 1.44 vs 0.43 in the most advantaged LGAs). What is particularly concerning is our finding that all-cause injury rates, and motor vehicle and low fall-specific injury rates, are increasing year-on-year at a greater rate in the most disadvantaged LGAs compared to the most advantaged LGAs. This indicates that inequities in injury are increasing, and urgent attention is required to address lower socioeconomic areas with increasing injury rates over time and/or relatively high injury rates. As our results relate to the socioeconomic level of the LGA, rather than the individual, these findings are suggestive of the role that more disadvantaged built environments play in an area being more unsafe.

The strengths of this study are the population-based capture of major trauma with known spatial coordinates of injury events, and the use of novel modelling approaches to quantify spatiotemporal variation in injury rates. Utilising spatiotemporal modelling, we were able to account for spatial correlation and temporal dependency, and for potentially sparse data, by borrowing strength from neighbouring areas. The methods that we have employed can be applied to other settings across the world to similarly inform the targeting of injury prevention interventions. However, a number of limitations exist. First, small counts across LGAs for specific causes of injury (with the exception of motor vehicle collisions and low falls) prevented a greater focus on these specific causes. Second, spatial aggregation of point data to areal data naturally leads to a loss in fidelity, and an inability to further our understanding on intra-LGA variability. Third, given that we explored LGA-specific socioeconomic level and not the socioeconomic level of the individual, there may be misclassification of individual-level socioeconomic status in situations where individuals are injured outside of their residential LGA; these findings are to be interpreted as area-level socioeconomic status only. Fourth, single measures of ARIA+ and IRSAD were used to classify remoteness and socioeconomic level, respectively. It is known that these measures vary within LGAs, and combined with the absence of individual-level socioeconomic level, these single measures are crude estimates. Finally, population-based incidence rates were calculated as exposure rates were not available for all causes of injury. In the absence of robust exposure measures, however, the underlying population often acts as the most appropriate denominator when exploring injury rates across large regions.^23^

## CONCLUSION

This study has demonstrated the power of spatiotemporal modelling in measuring the spatial and temporal variation in major trauma. These findings have important implications for injury prevention, by enabling the targeting of initiatives to areas with relatively, and significantly, high area-specific injury risk and/or areas with significantly increasing injury relative risk over time. Further, we demonstrated increasing rates of injury in the most disadvantaged areas, highlighting the need for a greater emphasis on reducing inequities in injury. Partnering with local, state and federal government and using these findings to target injury prevention initiatives is critical to reducing the burden of injury.

## Data Availability

All data produced may be requested through VSTORM: https://www.monash.edu/medicine/sphpm/vstorm/data-requests

## ACKNOWLEDGEMENTS

We thank the Victorian State Trauma Outcome Registry and Monitoring (VSTORM) group for providing VSTR data and Sue McLellan for her assistance with data preparation.

